# Disrupted PGR-B and ESR1 signaling underlies preconceptional defective decidualization linked to severe preeclampsia

**DOI:** 10.1101/2021.07.22.21260977

**Authors:** Tamara Garrido-Gomez, Nerea Castillo-Marco, Mónica Clemente-Ciscar, Teresa Cordero, Irene Muñoz-Blat, Alicia Amadoz, Jorge Jimenez-Almazan, Rogelio Monfort, Reyes Climent, Alfredo Perales, Carlos Simón

## Abstract

Decidualization of the uterine mucosa drives the maternal adaptation to invasion by the placenta. Appropriate depth of placental invasion is needed to support a healthy pregnancy; shallow invasion is associated with the development of severe preeclampsia (sPE). Maternal contribution to sPE through failed decidualization is an important determinant of placental phenotype. However, the molecular mechanism underlaying the *in vivo* defect linking decidualization to sPE is unknown. Here, we discover the footprint encoding this decidualization defect comprising of 166 genes using global gene expression profiling in decidua from women who developed sPE in a previous pregnancy. This signature allowed us to effectively segregate samples into sPE and control groups. Estrogen receptor 1 (*ESR1*) and progesterone receptor B (*PGR-B*) were found highly interconnected with the dynamic network of defective decidualization fingerprint. *ESR1* and *PGR-B* gene expression and protein abundance were remarkably disrupted in sPE. Thus, the transcriptomic signature of impaired decidualization implicates dysregulated hormonal signaling in the decidual endometria in women who developed sPE. These findings reveal a potential footprint that may be leverage for a preconception or early prenatal screening of sPE risk, thus improving prevention and early treatments.

## Introduction

Preeclampsia (PE) is a severe complication of late pregnancy and is the second leading cause of maternal mortality in the US, affecting 8% of first-time pregnancies (1). PE is characterized by the onset of hypertension, proteinuria, and other signs of maternal vascular damage that contributes to neonatal mortality and morbidity (1). Severe preeclampsia (sPE) is diagnosed based on elevated blood pressure (systolic ≥ 160 or diastolic of ≥ 100 mm Hg) or thrombocytopenia, impaired liver function, progressive renal insufficiency, pulmonary edema, or the onset of cerebral or visual disturbances (2). sPE is a placental insufficiency syndrome mediated by early deficient extravillous trophoblasts (EVTs) invasion of uterine decidua and spiral arterioles, leading to incomplete endovascular invasion and altered uteroplacental perfusion (3-5). Why shallow EVTs invasion occurs, however, remains to be determined (6).

Pregnancy health is dictated by the embryo, placenta, and the quality of the maternal decidua, where EVTs invasion and remodeling of maternal spiral arteries occur (7, 8). Accumulated evidence suggests that the contribution of the decidua to the etiology of PE (9), sPE (10-12), and placenta accreta (13) is significant, and cellular signaling in the decidua may determine whether these conditions develop. Decidualization is the remodelling of the endometrium initiated after ovulation necessary for adequate trophoblast invasion and subsequent placentation (14). Defective decidualization entails the inability of the endometrial compartment to undertake tissue differentiation, leading to aberrations in placentation and compromising pregnancy health (12).

In humans and other great apes, the formation of the decidua is a conceptus-independent process driven by progesterone and the second messenger cyclic adenosine monophosphate (15) that stimulates synthesis of a complex network of intracellular and secreted proteins through progesterone receptor activation. Endometrial decidualization involves secretory transformation of uterine glands (16), influx of specialized immune cells, vascular remodeling, and morphological (17, 18), biochemical (19, 20), and transcriptional reprogramming of the endometrial stromal compartment (21). We recently characterized the transcriptomics of human decidualization at single-cell resolution from secretory endometrial samples and showed that the process is initiated gradually after ovulation with a direct interplay between stromal fibroblasts and lymphocytes (21). However, most knowledge on decidual function in health and disease comes from *in vitro* model systems (22-24).

In the present study, we aimed to discern the preconception decidual transcriptomic signature associated with *in vivo* defective decidualization. We performed a comparative global transcriptional profiling of endometrium in women who developed sPE in a previous pregnancy. Initially, we identified 859 genes differentially expressed in sPE compared to control cases. Then, molecular DD-fingerprint of 166 genes associated with the development of sPE was defined and evaluated as a diagnostic tool in an independent cohort of samples. Finally, we identified progesterone receptor (PR) and estrogen receptor 1 (ER1) as targets of the DD-fingerprint genes. Our findings indicate that an endometrial transcriptomic signature persists years after the affected pregnancy. This signature may be leveraged for a preconception or early prenatal screening strategy in assessing sPE risk and may inform the development of sPE therapies.

## Results

### Endometrial transcriptome alterations during decidualization in sPE

We determined the transcriptional profiles of endometrial biopsies obtained in the late secretory phase in women who developed sPE in a previous pregnancy (n=24) and compared them to controls who had never had sPE (n=16). Controls included women who had a preterm birth with no signs of infection (n=8) and women who gave birth at full term with normal obstetric outcomes (n=8). Maternal clinical and neonatal characteristics of the participants are summarized in supplementary data, ***Supplementary file 1***. To rule out bias based on gestational age at delivery, preterm and term control specimens were compared. Principal component analysis (PCA) demonstrated an absence of clustering of samples based on gestational age (***Figure supplement 1A***). Volcano plotting showed no differentially expressed genes (DEGs) between preterm and term control samples (***Figure supplement 1B***).

Our experimental design randomly split samples into two cohorts, a training set (70%) and a test set (30%) (***Figure 1A***). Random sampling occurred within each class (sPE and controls), so overall class distribution of the data was preserved. The training set (n=29) was used for the identification of molecular fingerprinting encoding DD in sPE, while the test set (n=11) was used to confirm our findings. All samples in both cohorts were processed and RNA-sequencing performed in the same manner.

**Figure 1.**
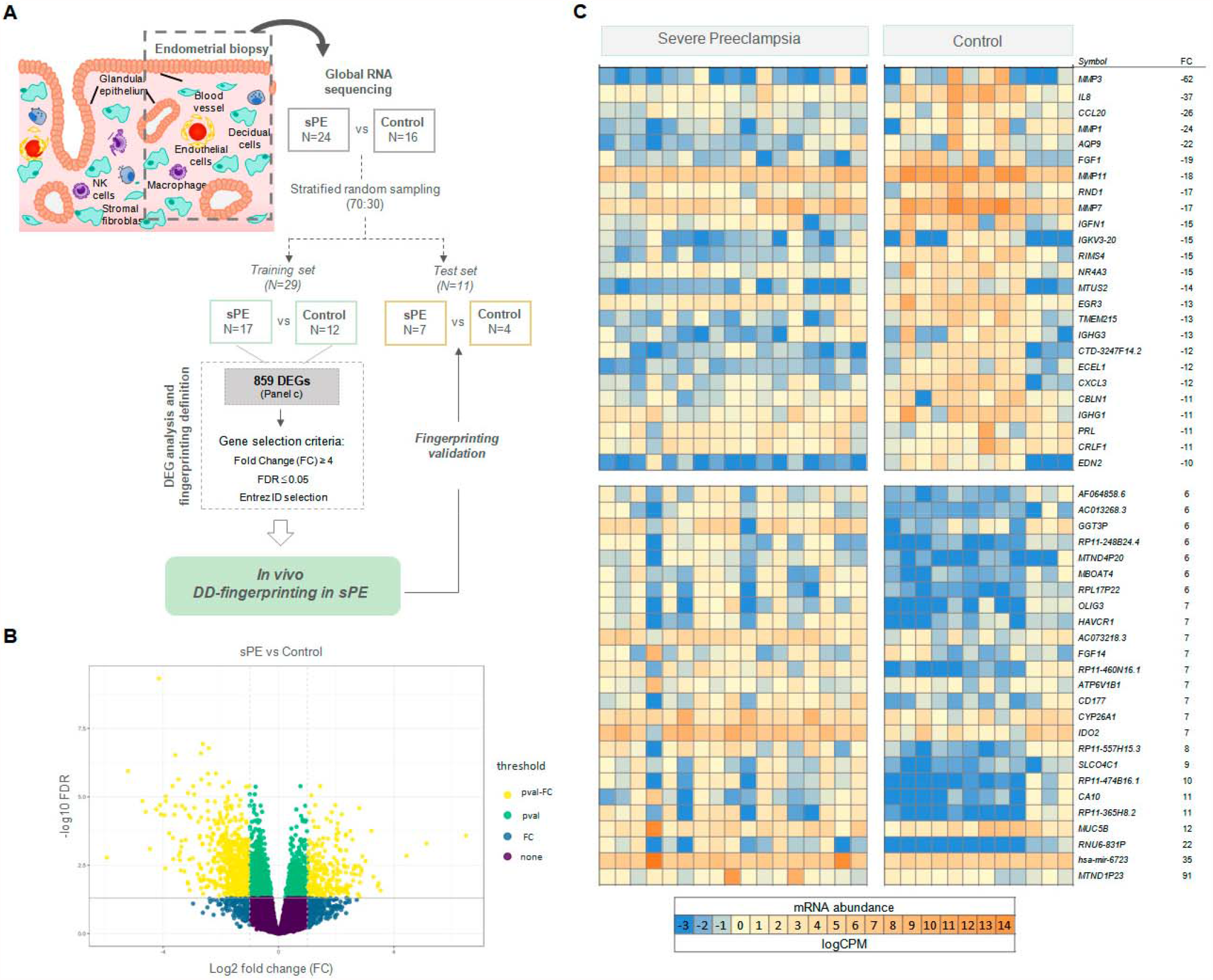
Global transcriptomics RNA-seq results revealed 859 DEGs in sPE vs control samples. (**A**) Schematic drawing of the study design used to identify and validate DD fingerprinting in sPE. (**B**) Statistical significance versus gene expression fold change is displayed as a volcano plot drawing the global RNA-seq results. Threshold indicates: pval-FC (FDR_≤_ 0.05 and log2-fold-change_≥_ 1); pval (green dots; FDR_≤_ 0.05 and log2-fold-change>1); FC (blue dots; FDR>0.05 and log2-fold-change_≥_ 1) and none (purple dots; FDR>0.05 and log2-fold-change>1). (**C**) Heatmap showing the 25 most upregulated and downregulated genes (total= 859; supplementaryx, Table 2) of control vs. sPE samples. **Supplementary File 1**. Maternal and neonatal characteristics for endometrial donors. **Supplementary File 2**. Differentially expressed genes in sPE vs control cases obtained from Global RNA-Seq analysis (859 DEGs). **Figure supplement 2**. Additional RT- qPCR validation.

Global RNA sequencing (RNA-seq) analysis in the training set was performed by comparing gene expression patterns in sPE (n=17) and controls (n=12). After quality trimming and filtering, reads were aligned to the reference genome hg19. Raw sequencing genes among the 29 samples numbered 56,638, and after normalization the number of genes included in the analysis was 18,301. Transcriptional analysis revealed 859 DEGs between sPE and controls (FDR ≤0.05 and FC≥ 2). The yellow dots shown in the volcano plot in Figure 1B denote statistically significant DEGs (***Figure 1B***). A total of 262 upregulated and 597 downregulated genes were differentially identified as being associated with DD in sPE (***Figure 1C***; complete list in ***Supplementary file 2***).

Downregulated transcripts include those involved in decidualization, such as *MMP3, PRL, IL-11, IHH*, and *SGK1*; and genes associated with signaling (e.g., *NR4A3* and *IL8*), growth factors (e.g., *FGF1* and *FGF14*), angiogenesis (e.g., *EDN2* and *TMEM215*), and immune response (*CCL20, CXCL3*, and *IGHG1*). Upregulated genes are involved in amino acid metabolic/catabolic processes (*GGT3P, IDO2*, and *PRODH*), transport, and oxidoreductase activity. We validated RNA-seq expression analysis by RT-qPCR in nine selected genes for sPE (n=14) compared to the control group (n=9) (***Figure supplement 2A***). Fold changes were highly concordant with the sequencing data (***Figure supplement 2B***).

### Comparison of DD transcriptomics in previous sPE *in vivo* vs *in vitro*

We previously described DD in human endometrial stromal cells (hESCs) isolated from women with previous sPE compared to women with normal obstetric outcomes, but this finding was restricted to the stromal cell population using an *in vitro* decidualization cell culture model (10). Here, we compared DD overlapping between DEGs reported *in vitro* (n=129) versus *in vivo* (n=859) in sPE compared to control women. Eighteen genes were differentially expressed between the two groups (FDR<0.05 and FC≥ 2) (***Figure 2A***); some genes were upregulated [e.g. *ERP27, CHODL*, and *PRUNE2*], and others genes were downregulated [e.g. *ISM1, MEST, MFAP2*, and *REEP2*] involved in decidualization failure in the sPE group. The expression pattern of common genes is presented as a box plot using counts per million, corroborating significant differential expression between sPE and control (***Figure 2B***). Expression was validated by RT-qPCR for five genes (***Figure 2C***), and high correlation (R=0.99) was observed with the global RNA-seq results (***Figure 2D***). Recently, *in vivo* transcriptomics of endometrium at single-cell resolution across the menstrual cycle was characterized (21). Transcriptome profile of stromal fibroblasts from late secretory phase allowed the identification of which deregulated genes in sPE from our RNAseq data are associated to hESC. We found that 300 genes from the 859 DEGs in sPE vs. control are expressed by hESC (***Figure 2E***). Taken together, the *in vivo* assessment provides a broad spectrum of dysregulated transcripts comparing with previous *in vitro* findings, that includes a high concordance with *in vivo* hESCs genes.

**Figure 2.**
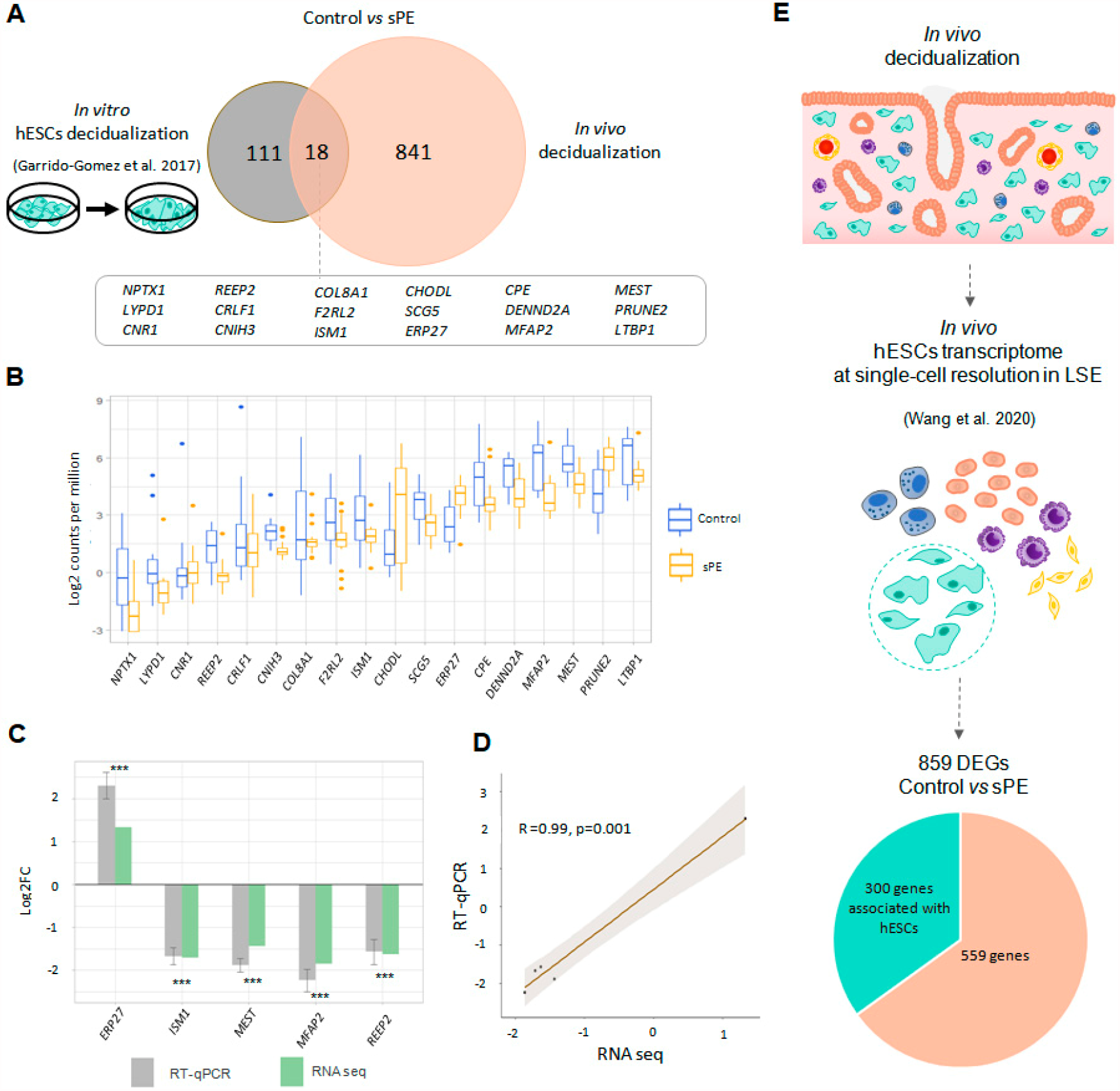
DD transcriptomics *in vitro* vs *in vivo*. **(A**) Common genes between previous *in vitro* (left) and current *in vivo* approaches to analyzing decidualization (right). Eighteen genes overlap in both approaches. (**B**) Box plot showing the average expression of the 18 common genes between control (blue boxes) and sPE (orange boxes) samples. **(C**) Fold change between control and sPE was validated by RT-qPCR (gray bars) and sequencing (green bars) for five transcripts from the 18 common transcripts identified in both approaches. (**D**) Correlation plot for RT-qPCR and RNA-seq (Pearson R = 0.99, p = 0.001). RT-qPCR values are expressed Mean± SE. *** p ≤ 0.001. (**E**) From the 859 DEGs obtained by global RNA sequencing, a subset of 300 DEGs were identified as genes with an hESCs origin using the scRNAseq data published by Wang et al. 2020.

### Identification of the fingerprint encoding human endometrial DD

To formulate the transcriptomic signature that encodes DD detected in sPE *in vivo*, we selected genes with significant dysregulation (FDR≤0.05) and at least four-fold increase (FC ≥ 4) between sPE and control with assigned EntrezID. A volcano plot shows 166 DEGs meeting these criteria included in the final DD signature (***Figure 3A***; complete list of genes is included in ***supplementary file 3***).

**Figure 3.**
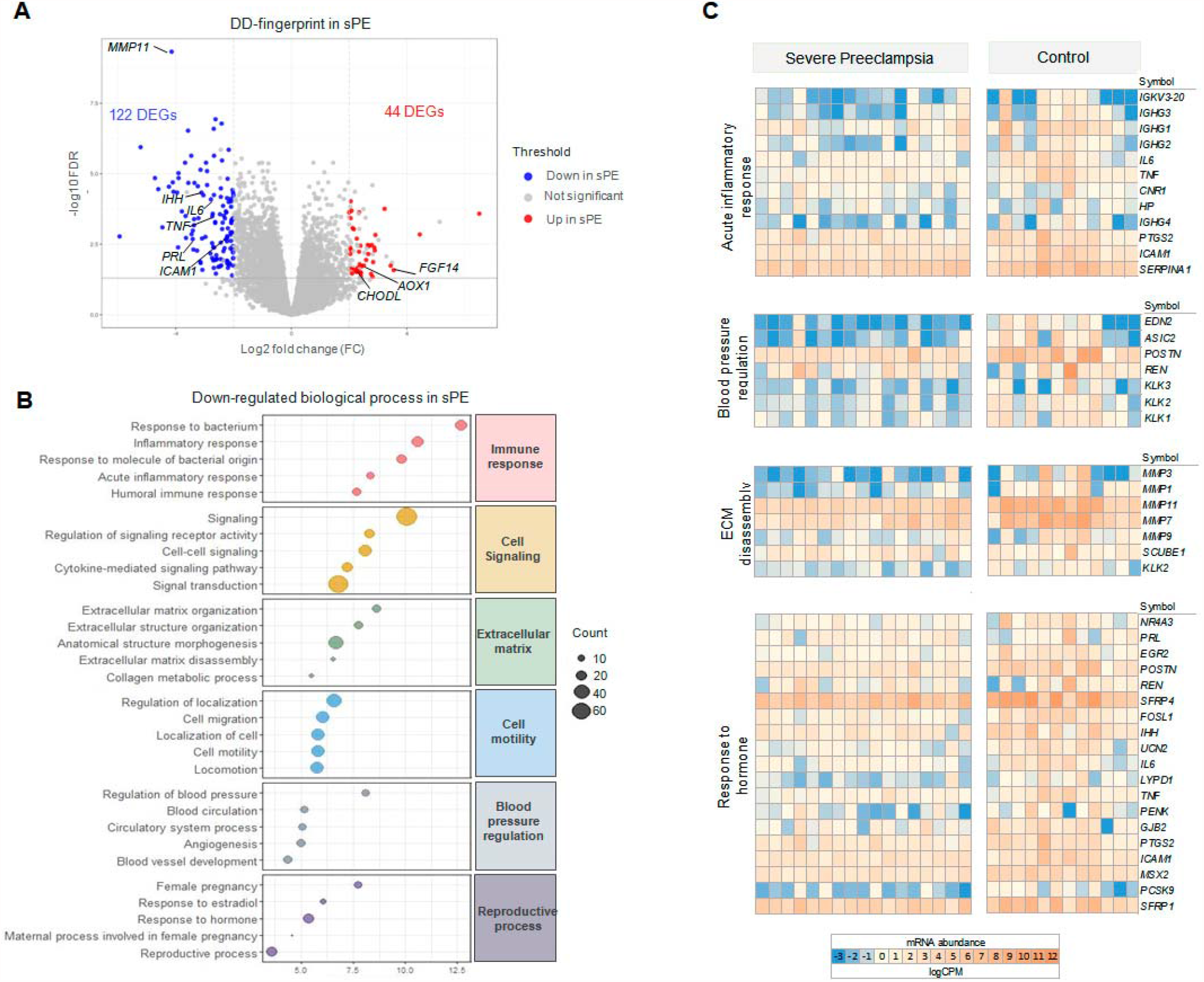
sPE-DD fingerprint composed of 166 DEGs. (**A**) Volcano plot showing downregulated (blue) and upregulated (red) genes in sPE from the DD-fingerprint. Each point represents one gene; gray points are the rest of genes obtained in the global RNA-seq analysis. (**B**) The five most highly downregulated biological process for each major category (*red*, immune response; *yellow*, cell signaling; *green*, extracellular matrix; *blue*, cell motility; *gray*, blood pressure regulation; *purple*, reproductive process). (**C**) Clustering of DD fingerprint genes shown for acute inflammatory response, extracellular matrix disassembly, regulation of systemic blood pressure, and response to hormones. **Supplementary file 3**. List of genes selected as defective decidualization signature in sPE (166 DEGs) **Supplementary file 4**. Biological process GO terms in sPE. **Figure supplement 3**. Additional 50 Biological Pathways.

Gene ontology analysis of the gene signature associated with DD in sPE identified 479 enriched biological processes (p < 0.005; ***supplementary file 4***). Downregulated pathways were associated with immune response, cell signaling, extracellular matrix, cell motility, blood pressure regulation, and reproductive processes (***Figure 3B***). All are hallmarks of impaired decidualization and sPE pathogenesis. The 50 most enriched pathways are presented in ***Figure supplement 3***. We identified fingerprinting genes representative of the altered pathways in sPE, such as *IL6* and *TNF*, regulating the acute inflammatory response, *MMP3* and *MMP1* participating in the extracellular matrix disassembly, *POSTN* and *REN* as affected regulators of systemic blood pressure, and *PRL, IHH*, and *ICAM1*, implicated in the downregulated response to hormones (***Figure 3C***). Upregulated biological processes were associated with metabolism, the nervous system, interaction between organisms, and negative regulation of chorionic trophoblast cell proliferation. Functional analysis revealed that the 166 DEGs included in DD fingerprinting are implicated in pathways related to decidualization, corroborating the maternal contribution to sPE. Interestingly, the number of downregulated genes was higher than the number of upregulated genes in sPE compared to controls, suggesting that, *in vivo*, DD may be induced by the lack of expression of a subset of genes.

Based on the 166 genes included in the DD signature, PCA showed that sPE and control samples clustered separately in two groups, except for three control samples (C20, C21, and C22) (***Figure 4A***). High variance between groups was effectively captured in the first two principal components. Unsupervised hierarchical clustering analysis confirmed that gene fingerprinting effectively segregated the two groups: one encompassing mainly controls and the other mainly sPE samples (***Figure 4B***). The same three controls clustered with the sPE group, recreating the PCA results.

**Figure 4.**
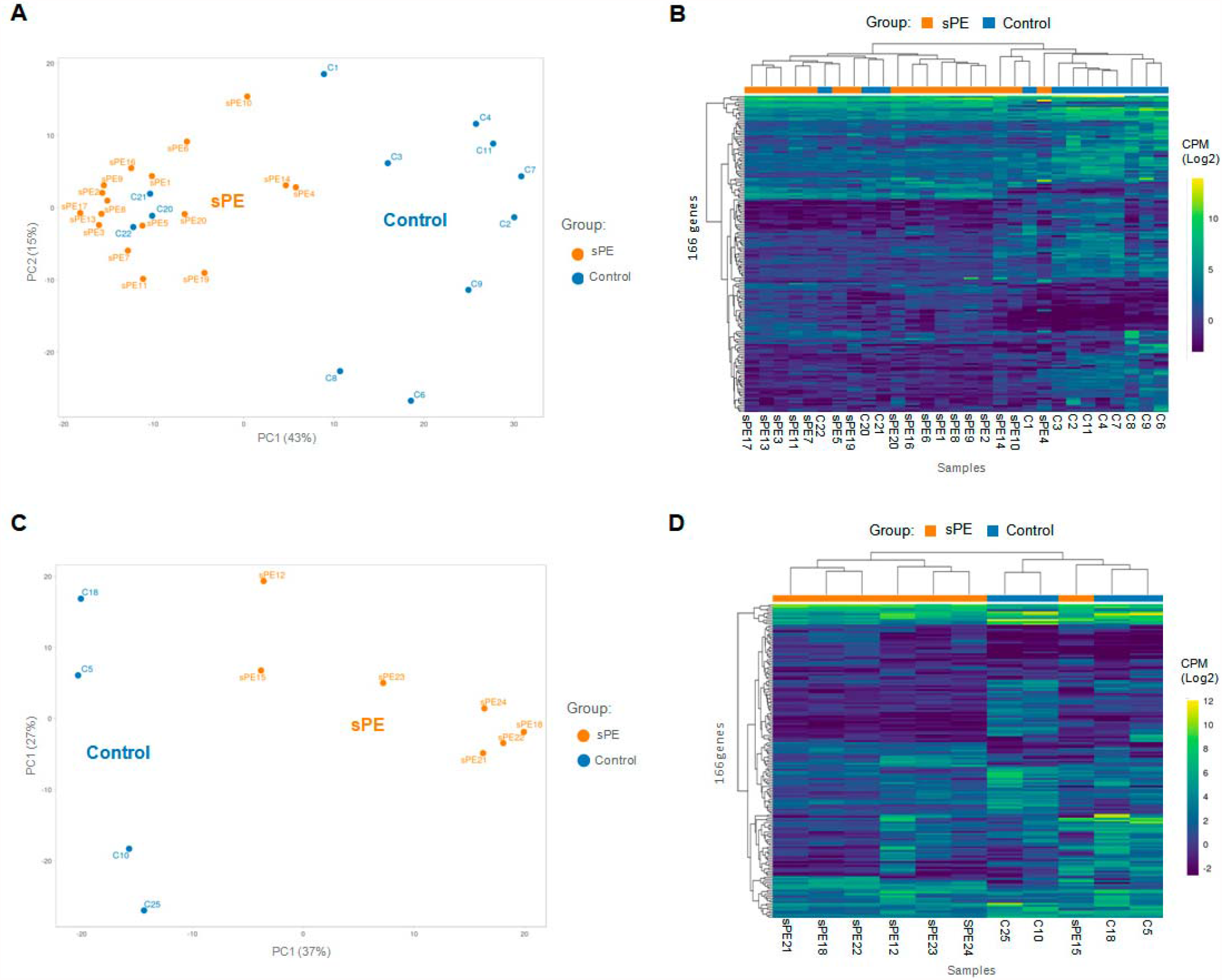
Validation of the DD fingerprint in Spe. (**A**) PCA based on 166 genes included in the fingerprinting in the training set. Each sample is represented as a colored point (*blue*, control; *orange*, sPE). (**B**) Heatmap dendogram of expression of the 166 genes included in the final fingerprinting for each sample of the training set (control, n=12; sPE, n=17). (**C**) PCA based on the fingerprinting in the test set. Each sample is represented as a colored point (*blue*, control; *orange*, sPE). (**D**) Heatmap dendogram of expression of the 166 genes included in the final fingerprinting for each sample of the test set (control, n=4; sPE, n=7).

To validate the DD gene signature in an independent cohort of samples [sPE (n=7) vs. control (n=4)], PCA based on these transcripts effectively segregated samples in two homogeneous groups (***Figure 4C***), corroborated by hierarchical clustering (***Figure 4D***). These genes successfully grouped 100% of controls and 85.7% of sPE cases supporting DD in sPE.

### DD fingerprint in sPE is connected to ER1 and PR-B

Of the 166 genes in the DD signature, 106 endometrial enriched genes encode for specific proteins reported by the Human Protein Atlas (25). Interestingly, 37 of those genes (35%) were included in the transcriptome modulated by *ESR1* (26), and 54 genes (51%) overlapped with the transcriptome and cistrome associated with *PGR* (27) (***Figure 5A***).

**Figure 5.**
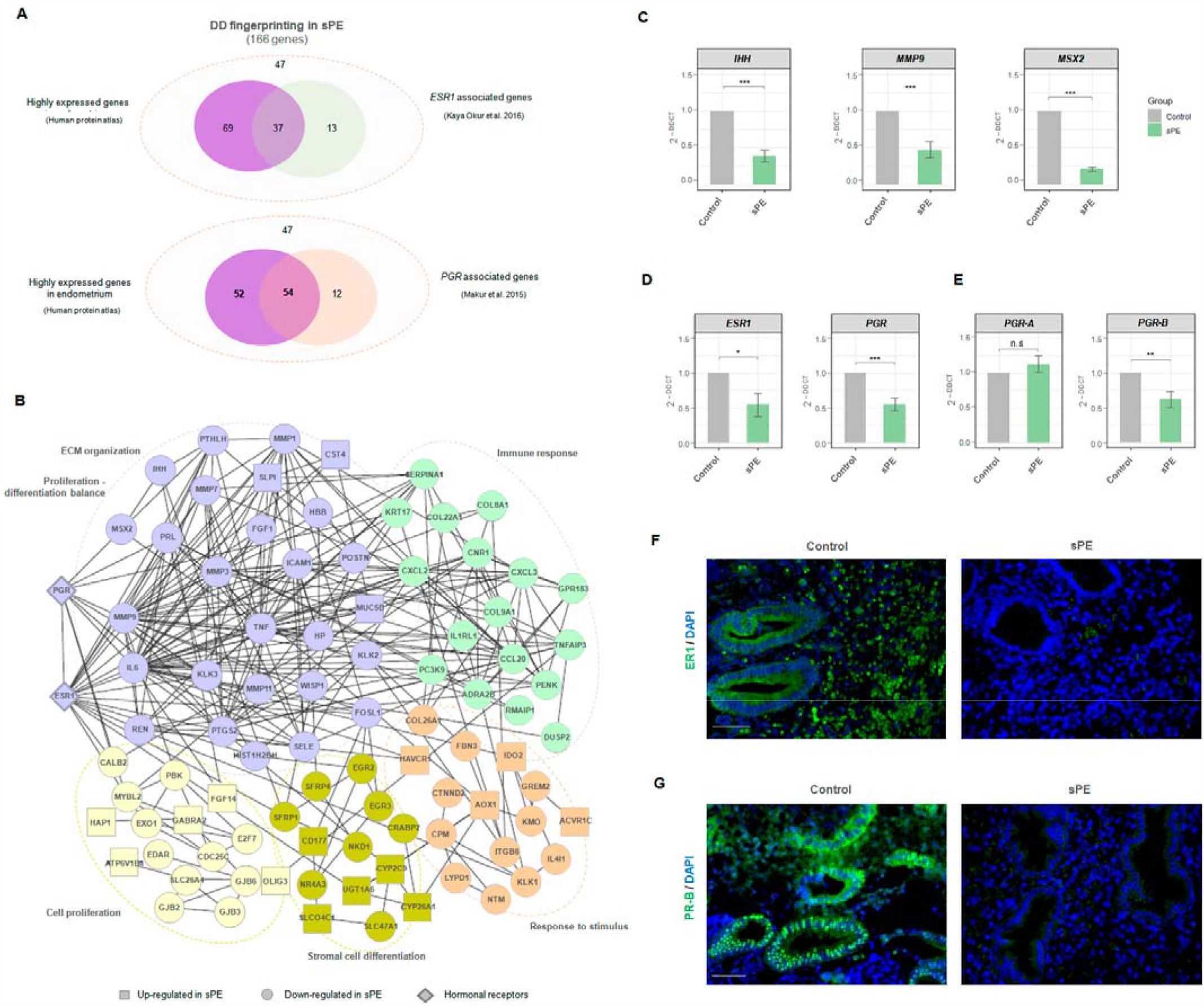
ER1 and PR-B are linked to DD fingerprinting in sPE. (**A**) Venn diagram displaying genes included in the fingerprinting (166) predominantly expressed in the endometrium based on human protein atlas data that overlap with genes modulated by *ESR1* described by Okur et al., 2016 (53) and genes associated with *PGR* silencing described by Mazur et al., 2015 (27). (**B**) Network showing the connections between proteins codified by DD fingerprinting and the hormonal receptors, ER1 and PR. *Squares*, significantly up-regulated genes; *circles*, significantly downregulated genes; *rhombus*, ER1 and PR. Colors indicate different clusters established by String k-means method. (**C-E**) Gene expression levels of *IHH, MMP9, MSX2, ESR1, PGR, PGR-A*, and *PGR-B* assessed for sPE (n= 13) vs. controls (n=9) by RT-qPCR (*gray bars*, control; *green bars*, sPE). RT-qPCR values are expressed as mean± SE. *** p<0.001, ** p<0.01, *p<0.05. (**F-G**) Tissue sections of control (n=4) and sPE (n=4) endometrium during late secretory phase were immunostained with antibody against ER1 or PR. Nuclei were visualized with DAPI. Scale bar: 50 µM.

We evaluated the interaction between steroid receptor signaling and the proteins encoded by DD-fingerprinting genes in sPE by building a dynamic network including ER1 and PR. String software (28) was used to construct network connections visualized with Cytoscape software (29). The interactome contained 105 nodes directly interconnected by 322 edges (***Figure 5B***). This DD fingerprint network showed a highly enriched protein–protein interaction (PPI) in sPE; indeed, the interconnection between nodes was significantly higher than the 119 edges expected (PPI enrichment p < 1.0e-16). Clustering revealed five main modules based on their connectivity degree, with functionally relevant genes involved in the regulation of proliferation–differentiation balance, response to stimulus, immune response, and extracellular matrix organization. Interestingly, both ER1 and PR were strongly embedded in the network and highly connected with DD fingerprinting highlighting the interaction of hormonal receptors with notable decidualization mediators such as *IHH, MMP9*, and *MSX2* validated by RT-qPCR (***Figure 5C***). Furthermore, the interactome demonstrated a direct interaction between ER1 and PR. These results support the transcriptomic dysfunction of the genes present in the DD signature through imbalanced hormone receptor signaling in sPE.

We then analyzed the expression of *ESR1* and *PGR* in the endometrial tissue from a subset of women with prior sPE (N=13) compared to controls (N=9) by RT-qPCR. We found reduced expression of transcripts encoding the hormone receptors *ESR1* (p < 0.05) and *PGR* (p < 0.001) in sPE patients (***Figure 5D***). In depth expression analyses revealed that the isoform *PGR-B* was significantly downregulated in sPE vs controls (p < 0.01), while the isoform *PGR-A* was unaffected (p > 0.05) (***Figure 5E***). These results were confirmed at the protein level by immunohistochemical analysis of ER1 and PR-B in endometrial biopsies collected in the late secretory phase from women with previous sPE (N=4) and controls (N=4) (***Figure 5F and 5G***). Both receptors were highly expressed through the decidualized endometrium, especially in the secretory glands in controls. In contrast, their expression was greatly reduced or absent in sPE samples. These results suggest that the DD transcriptomic signature implicates dysregulated ER1 and PR-B signaling in the late secretory phase in sPE patients.

## Discussion

Most scientific and clinical diagnostic efforts in sPE focus on placental surrogates. In this context, shallow cytotrophoblast invasion induces deficient vascular remodeling and ultimately aberrant placentation. This leads to placental ischemia and the release of soluble factors that induce the maternal syndrome, including the disbalanced levels of soluble fms-like tyrosine kinase 1 (sFLT1) and placental growth factor (PlGF) (30, 31). sFLT1 protein binds to PlGF, preventing its interaction with endothelial receptors and leading to endothelial dysfunction. Accordingly, sFLT1 is increased while free PlGF is decreased in serum from women with PE (32). The sFLT1/PlGF ratio has been proposed as a biomarker showing a positive predictive value of 36.7%, with 66.2% sensitivity and 83.1% specificity but its application is effective only four weeks before PE symptoms manifest (33). Therefore, during the first trimester there is a lack of highly sensitivity and specificity screening methods to detect sPE early and prevent mortality and morbidity.

Current strategies based on placental dysfunction provide delayed results for preventive interventions and new approaches are urgently needed. Recent studies suggest that sPE might also be a disorder of the decidua, opening new avenues (9, 10, 34). In this sense, our previous work demonstrated that hESC isolated from patients with previous sPE failed to decidualize *in vitro*, suggesting a role of the maternal factor in the development of this disease (10).

In the present study, we highlighted the underlying molecular defect that may explains *in vivo* decidualization failure as important contributor to shallow placental invasion in sPE. For this purpose, we investigated the role of the decidua by leveraging global RNA-seq in late secretory endometrium. We identified 859 genes differentially expressed in sPE compared to controls, including genes involved in decidualization such as *PRL, IL6*, and *IHH*, as well as several novel genes. Eighteen of the differentially expressed genes overlapped with DEGs obtained in our previous *in vitro* decidualization study. *In vivo* endometrial biopsies include other cells in addition to hESCs, and thus the expected degree of overlap between both *in vitro* and *in vivo* approaches should be modest as such was observed. However, we identified a large percentage of those DEGs that were associated with *in vivo* transcriptomic profile of hESCs resolved at cell-level from a healthy late secretory endometrium. Thus, the high number of our identified *in vivo* DEGs reflects the high complexity of decidualization at cellular heterogeneity in a physiological stage of women with previous sPE. From these, we identified a DD signature composed of 166 genes associated with sPE.

This sPE-DD signature includes genes that allowed us to segregate samples from the training set into sPE and control groups, which were confirmed by the test set. Only three control specimens were misclustered in the training set and one sPE was misclustered in the test set, such that 90.9% of samples from an independent cohort were properly clustered in the dendogram. Decidualization is a highly dynamic process governed by: (i) inter-individual variability in the endometrial menstrual cycle supported by displacement of the window of implantation in one out for patients suffering from recurrent implantation failure (21, 35), (ii) the physiology of the spatial expansion of decidualization process, starting in some areas around spiral arteries and extending to the entire endometrium during the last days of the menstrual cycle (14), (iii) and the random spatial sampling during the endometrial biopsy that could influences cell types proportions, which is inherent to the experimental strategy used in our investigation. Decidua sample segregation is consistent with the results presented by Munchel et al. (36), who classified PE patients based on circulating RNA (C-RNA). In our study, we determined that the gene expression associated with DD had the highest potential to segregate sPE.

Interestingly, most of the DD-fingerprint genes in sPE were related to the downregulation of *ESR1* and *PGR*, specifically *PGR-B*. We hypothesize that low expression of *PGR-B* and *ESR1* activate endometrial decidualization by dysregulating progesterone (P4) and estrogen (E2) action. Consequently, P4 and E2 related cellular signaling may be compromised, leading to the development of DD. Stromal cell differentiation, stromal–epithelial crosstalk (37), extracellular matrix degradation (38), immune system response, and endothelial function (39) may all be disrupted by low expression of *PGR-B* and *ESR1*. Remarkably, the local immune system appeared to be dysregulated due to the altered expression of interleukins, cytokines, chemokines, and immunoglobulin, which could disrupt the tolerant microenvironment at the maternal-fetal interface in pregnancy and contribute to the development of sPE (40, 41).

Based on our findings, we postulate that in non-sPE pregnancies, balanced hormonal signaling leads to proper decidualization, which in turn interacts with immune and endothelial cells to control cytotrophoblast invasion (***Figure 6A***). P4 and E2 activate their receptors in the epithelium and signal to the stromal compartment. Likewise, stromal PR is activated by ER1, which induces target genes involved in decidualization such as *MSX2, CNR1*, and *MMP9*. In contrast, in sPE, endometrial *ESR1* and *IHH* are downregulated, decreasing *PGR* expression (***Figure 6B***) and leading to compromised decidualization, endothelial dysfunction, and local immune dysregulation.

**Figure 6.**
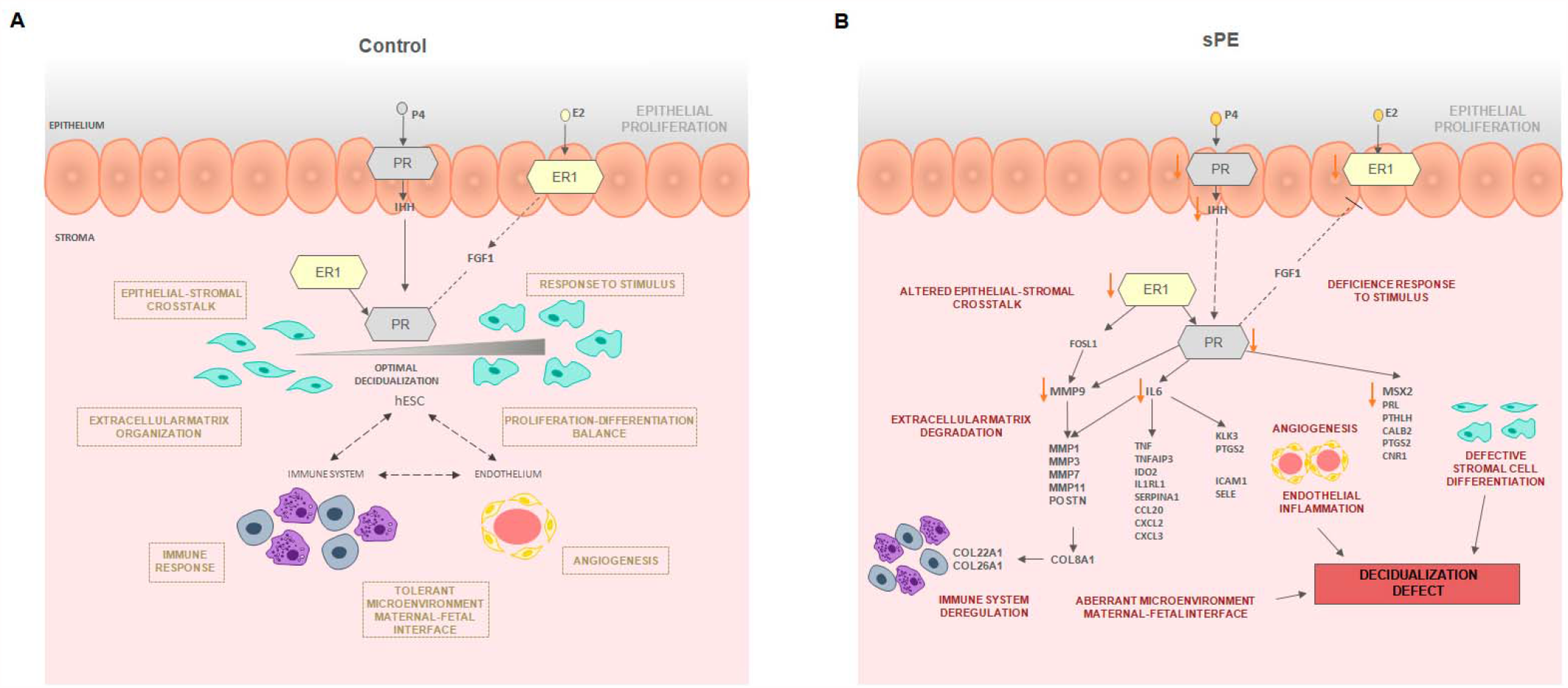
Modeling of the molecular mechanism for DD in sPE. (**A)** Decidualization induced by P4 and E2 in control pregnancy including the interaction of immune response and endothelium. **(B)** Hypothetical network that could link DD and dysregulated hormone signaling in sPE. All genes were downregulated. Biological processes specified are candidates to be impaired based on functions associated with the observed dysregulation. *Red arrows* show the downregulation of decidualization modulators.

In sPE, we found evidence of impaired epithelial-stromal crosstalk through *IHH* and *FGF1*, which could lead to an imbalance of hESC proliferation and differentiation (8, 42). In support of this, we observed downregulated genes involved in stromal cell differentiation including *PRL, CNR1, MSX2*, and *PTSG2*. Further, metalloproteinases such as *MMP9, MMP1, MMP7* and *MMP11* were downregulated in sPE, that could be associated to defective cell motility and extracellular matrix degradation. *MMP9* is regulated by both ER1 and PR and its downregulation is associated with impaired decidualization (43) and PE (44). Other transcriptomic deregulations in sPE are connected to endothelial and immune system dysfunction, such as *KLK3* and *PTGS2*, both of which are associated with altered angiogenesis (45, 46); *ICAM1* and *SELE* are involved in endothelial inflammation disbalance (47, 48); and *TNF* influences the tolerant microenvironment at the maternal-fetal interface (49). Taken together, our findings reveal a DD transcriptomic fingerprint in sPE driven by an imbalance in ER1 and PR signalling.

Our findings reveal significant gene expression dysregulation underlying DD in the late secretory phase in women who have had sPE. The potential origin of sPE may lie in the downregulation of *ESR1* and *PGR-B*. Both receptors are strongly coordinated to regulate decidualization and *PGR* expression is induced by ER1, which is inhibited by PR (50). E2 and P4 act through ER1 and PR in the epithelium and stroma and modulate the transcriptome and to promote crosstalk between both compartments (51). ER1 in the epithelium regulates stromal decidualization via paracrine mechanisms mediated by leukemia inhibitory factor (LIF), which controls *IHH* expression that transduce the signal activation of PR in the stroma (52). Also, stromal ER1 activates PR in the same compartment (53). In addition, both E2 and P4 modulate common targets, that are important markers that increases their expression during proper decidualization [e.g. *MMP9, PRL, IL6*]. These findings could inform the development of therapeutic targets to restore optimal decidualization in sPE. Further studies are needed to fully elucidate the hormone signaling pathways that become dysregulated in sPE and the role of decidualization resistance in the manifestation of the condition.

In conclusion, our findings reinforce a maternal cause for sPE through DD. This condition may result from an aberrant response to progesterone and estrogen mediated by ER1 and PR-B signaling. DD fingerprinting may allow for preconception and early prenatal sPE screening and could aid in the development of therapeutic strategies to treat this enigmatic pathological condition. Future studies should focus on the translational potential of the DD fingerprinting to develop new noninvasive strategies based on circulating RNA to improve diagnosis and prognostication for women with sPE.

## Materials and Methods

### Study design

A total of 40 non-pregnant women who experienced a previous pregnancy were enrolled in this study for endometrial RNA-sequencing analysis. Endometrial samples were obtained during late secretory phase in 24 women who had developed sPE in a previous pregnancy and in 16 women with no history of sPE with full term (n=8) and preterm pregnancies (n=8) as controls. Endometrial biopsies were processed to obtain RNA and then converted to cDNA for library generation to perform next generation sequencing. The experimental design was based on a stratified random sampling with a 70:30 proportion in two cohorts: a training (n=29) and validation (n=11) set of samples. The training set of samples was analyzed by RNA-seq to identify the global transcriptomic profiling changes between control (n=12) and sPE (n=17) samples. Selection criteria were applied to define a transcriptomic fingerprinting associated with DD detected in sPE. Finally, targeted analysis of the DD signature was validated in the test set composed of controls (n=4) and sPE (n=7).

### Human donors

Endometrial samples were collected from women aged 18–42 without any medical condition who had been pregnant 1–8 years earlier. All participants had regular menstrual cycles (26-32 days) with no underlying gynecological pathologic conditions and had not received hormonal therapy in the 3 months preceding sample collection. After the inclusion criteria were applied, endometrial biopsies were obtained by pipelle catheter (Genetics Hamont-Achel, Belgium) under sterile conditions in the late secretory phase (cycle days 22-32). Specimens were kept in preservation solution until processing. Maternal and neonatal characteristics of women with sPE and controls are summarized in ***supplementary file 1***. This study was approved by the Clinical Research Ethics Committee of Hospital La Fe (Valencia, Spain) (2011/0383), and written informed consent was obtained from all participants before tissue collection and all samples were anonymized.

### RNA extraction

Total RNA from endometrial biopsies was isolated using QIAsymphony RNA kit (Qiagen, Hilden, Germany) following the manufacturer’s protocol. RNA concentrations were quantified using a Multiskan GO spectrophotometer (Thermo Fisher Scientific, Waltham, US) at a wavelength of 260 nm. Integrity of the total RNA samples was evaluated using an Agilent 4200 TapeStation system (Agilent Technologies Inc., Santa Clara, CA) and the samples with RNA integrity were used for the global RNA-seq.

### Global RNA-seq library preparation and transcriptome sequencing

cDNA libraries from total RNA samples (n=40) were prepared by an Illumina TruSeq Stranded mRNA sample prep kit (Illumina, San Diego, CA). Three micrograms of total RNA were used as the RNA input according to the manufacturer’s protocol. mRNAs were isolated from the total RNAs by purifying the poly-A containing molecules using poly-T oligo attached to magnetic beads. The RNA fragmentation, first and second strand cDNA syntheses, end repair, single ‘A’ base addition, adaptor ligation, and PCR amplification were performed according to the manufacturer’s protocol. The average size of the cDNA libraries was approximately 350 bp (including the adapters). cDNA libraries were validated for RNA integrity and quantity using an Agilent 4200 TapeStation system (Agilent Technologies Inc., Santa Clara, CA) before pooling the libraries. The pool concentration was quantified by qPCR using the KAPA Library Quantification Kit (Kapa Biosystems Inc.) before sequencing in a NextSeq 500/550 cartridge of 150 cycles (Illumina, San Diego, CA). Indexed and pooled samples were sequenced 150-bp paired-end reads by on the Illumina NextSeq 500/550 platform according to the Illumina protocol.

### Quality control and pre-processing data

Once sequencing was completed, we checked for outliers or any technical issue in the raw data (sequence quality, read alignment, quantification, and reproducibility). After the initial quality assessment, pre-processing analyses were performed over the read counts data (low-count filter and normalization). Transcriptomic data were deposited in the Gene Expression Omnibus database (accession number GSE172381).

### Differential expression analysis

All sequences were pre-processed, normalized, and analyzed comparing sPE (n=17) to controls (n=12) from training set. We applied the trimmed mean of M-values normalization strategy to our gene expression values (54). The Bioconductor package edgeR (54) in R software was used. The p-value adjustment method was FDR with a cut-off of 0.05 and the minimum absolute log2-fold-change required was 1 (FC ≥ 2). A volcano plot was created to visualize DEGs. For a better overview, we distinguished between DEGs with a high or low fold-change. In this case, the threshold at the legend indicates: none (do not have DEGs); p-value (DEGs, but low fold-change: smaller than 1 in absolute value); FC (high fold-change, but not DEGs) and p-value–FC (both DEGs and high fold-change: log2-fold-change greater than 1 in absolute value).

### Transcriptomic fingerprinting definition and validation

Genes with assigned EntrezID with an FDR cut-off of 0.05 and an expression ≥ 4-fold higher in the sPE vs. control training set samples were selected to define a fingerprint associated with DD in sPE. Targeted analysis of fingerprinting genes was performed using the validation set of samples. PCA and unsupervised hierarchical clustering with a Camberra distance based on gene signature were performed comparing sPE to control specimens.

### Enrichment analysis

Biological processes in which DEGs are involved were studied. In edgeR, GO analyses can be conducted using *goana*. An FDR cutoff of 5% is used when extracting DE genes and for logFC, we used a cut-off value of 1 [UP, logFC>1 and DOWN; logFC<(−1)]. The ontology domain that GO term belongs to is biological process (BP). Because the p-values obtained are not adjusted for multiple testing, we ignored GO terms with p-values greater than about 0.005.

### Interaction network

An interaction network between proteins encoded by DD fingerprinting genes was created using the functional analysis suite String (28). To construct the network, the interactions included were from curated databases and included experimentally determined and predicted interactions, text mining, co-expression information, and protein homology. The clustering algorithm k-means was applied based on the distance matrix obtained from the String global scores. The network was visualized using Cytoscape software (29).

### qRT-PCR gene validation

To validate our transcriptomic results, a selection of differentially expressed genes was validated by qRT-PCR in a subgroup of samples from the experimental cohort [controls (n=9) and sPE (n=14)]. Specific primers for each gene are described in ***supplementary file 5***. cDNA was generated from 400 ng of RNA using the SuperScript VILO cDNA Synthesis Kit (Thermo Fisher Scientific, Waltham, US). Template cDNA was diluted 5 in 20 and 1 µL was used in each PCR. Real-time PCR was performed in duplicate in 10 µL using commercially validated Kapa SYBR fast qPCR kit (Kapa biosystems Inc, Basilea, Switzerland) and the Lightcycler 480 (Roche Molecular Systems, Inc, Pleasanton, CA) detection system. Samples were run in duplicate along with appropriate controls (i.e., no template, no RT). Cycling conditions were as follows: 95°C for 3 min, 40 cycles of 95°C for 10 s, 60°C for 20 s, and 72°C for 1 s. A melting curve was done following the product specifications. Data were analyzed using the comparative Ct method (2−ΔΔCT). Data were normalized to the housekeeping gene β-actin, changes in gene expression were calculated using the ΔΔCT method with the control group used as the calibrator; values are illustrated relative to median in the control group. The relative expression of *PGR-A* mRNA was calculated by subtracting the relative expression of *PGR-B* mRNA from that of *PGR* total.

### Immunofluorescence of tissue sections

Endometrial tissue samples were fixed in 4% paraformaldehyde and preserved in paraffin-embedded blocks. For immunostaining, tissue sections were deparaffinated and rehydrated. Antigen retrieval was performed with buffer citrate 1x at 100°C for 10 min. Then, non-specific reactivity was blocked by incubation in 5% BSA/0.1% PBS-Tween 20 at room temperature for 30 min. Sections were incubated at room temperature for 1.5 h with primary antibodies (1:50 rabbit monoclonal anti-human progesterone receptor, Abcam, Cambridge, UK) and 1:50 mouse monoclonal anti-human estrogen receptor 1 (Santa Cruz Biotechnology, CA, USA) diluted in 3% BSA/0.1% PBS-Tween 20. Then, slides were washed two times for 10 min with 0.1% PBS-Tween 20 before they were incubated for 1 h at room temperature with AlexaFluor-conjugated secondary antibodies diluted in 3% BSA/0.1% PBS-Tween 20. Finally, slides were washed two times in 0.1%PBS-Tween 20. To visualize nuclei, 4′,6-diamidino-2-phenylindole at 400 ng/uL was used. Tissue sections were examined using a EVOS M5000 microscope.

### Statistical analysis

Clinical data are expressed as mean ± standard error mean (SEM). Clinical data were evaluated by Student’s t-test for comparisons between sPE and control samples. Statistical significance was set at p ≤0.05. Differential expression analysis was performed using the R package edgeR.

## Data Availability

Transcriptomic data were deposited in the Gene Expression Omnibus database (accession number GSE172381).

## Acknowledgements

We are grateful to Dr. Alfredo Perales and the Obstetric Area from the Hospital La Fe for invaluable help in the enrollment of participants and obtaining the tissue samples that made this study possible. We thank the Hospital La Fe recruiters Rogelio Monfort, Reyes Climent, Laura Rubert, Joana Dasí, and Julia Escrig for their assistance in compiling clinical data. We are indebted to the patient participants. This work was supported by the grant PI19/01659 from the Spanish Carlos III Institute awarded to T.G.-G. N Castillo-Marco was supported by the PhD program FDGENT/2019/008 from the Spanish Generalitat Valenciana. I Muñoz-Blat was supported by the PhD program PRE2019-090770 and funding was provided by the grant RTI2018-094946-B-100 from the Spanish Ministry of Science and Innovation with C Simón as principal investigator. This work was funded partially by Igenomix S.L.

## Author contributors

T.G.-G., N.C.-M., I.M.-B. and T. C. performed the experiments. R.M., R.C. and A.P. enrolled study participants and collected endometrial samples. M.C., A.A., J.J. and N.C.-M. generated transcriptional profiling data. T. G.-G., A.P., and C.S. designed the experimental plan and guided its execution. All authors contributed to the interpretation of data. T.G-G., N.C.-M., I.M.-B., and T. C. prepared the figures and tables. T.G.-G., N.C.-M., M.C., and C.S. wrote the manuscript, which the other authors helped edit.

