## Supplementary file 1 for "Disrupted PGR-B and ESR1 signaling underlies preconceptional defective decidualization linked to severe preeclampsia"

**Supplementary Table 1.** Maternal and neonatal characteristics for endometrial donors.

|  | **sPE (n=24)** | **Control** | |  |  |  |
| --- | --- | --- | --- | --- | --- | --- |
|  |  | **Term pregnancy (n=8)** | **Preterm pregnancy (n=8)** |  | **P* (sPE vs term)** | **P* (sPE vs preterm)** |
| Maternal age (years) | 37.3 (0.8) | 37.6 (1.1) | 34.9 (2.4) |  | n.s. | n.s. |
| Systolic blood pressure (mm Hg) | 164.8 (3.5) | 124.0 (6.6) | 113.1 (7.7) |  | < 0.001 | < 0.001 |
| Diastolic blood pressure (mm Hg) | 99.0 (2.1) | 69.4 (2.4) | 62.8 (3.0) |  | < 0.001 | < 0.001 |
| Proteinuria (mg/dL) | 280.2 (42.5) | 0 or NA | 0 or NA |  | NA | NA |
| Gestational age at delivery (weeks) | 31.9 (0.6) | 39.5 (0.4) | 33.9 (1.2) |  | < 0.001 | n.s. |
| Birth weight (g) | 1651.6 (153.3) | 3200.1 (160.3) | 2403.8 (218.3) |  | < 0.001 | < 0.05 |
| Parity (n) | 1.7 (0.2) | 3.4 (0.7) | 2.8 (0.7) |  | < 0.01 | < 0.05 |
| Interval from last pregnancy to endometrial biopsy (years) | 3.4 (0.3) | 4.7 (1.0) | 3.3 (1.0) |  | n.s. | n.s. |
| Mean ± SEM * One-tailed student´s t-test |  |  |  |  |  |  |
| NA: Not available |  |  |  |  |  |  |
| n.s.: Not significant |  |  |  |  |  |  |
