## Supplementary file 5 for "Disrupted PGR-B and ESR1 signaling underlies preconceptional defective decidualization linked to severe preeclampsia"

**Supplementary Table 5.** RT-qPCR primers list.

| **Sequence Name** | **Sequence** |
| --- | --- |
| AOX1_FW | **TGTCCATCTACACGCTGCTC** |
| AOX1_RV | **TCCTCAAATTCTGGCAATCC** |
| ERP27_FW | **ACAAGGCCTCCCCAGAGTAT** |
| ERP27_RV | **CTTCTGCTGTGGGCAGTGTA** |
| ISM1_FW | **GACCTGTGACCGTCCAAACT** |
| ISM1_RV | **AGAACTCGCTTTTGCAGCTC** |
| MEST_FW | **CGCAGGATCAACCTTCTTTC** |
| MEST_RV | **CATCAGTCGTGTGAGGATGG** |
| MFAP2_FW | **CCAGATCGACAACCCAGACT** |
| MFAP2_RV | **GCAAGGCCTGTGTATGGAGT** |
| MMP11_FW | **GGTCTCTGAGGGTCAAGCAG** |
| MMP11_RV | **AGTTCATGAGCTGCAACACG** |
| PGRMC1_FW | **CCTCTGCATCTTCCTGCTCT** |
| PGRMC1_RV | **CGTTGATGGCCATGAGTATG** |
| PGR_FW | **GTGGGAGCTGTAAGGTCTTCTTTAA** |
| PGR_RV | **AACGATGCAGTCATTTCTTCCA** |
| PGRB_FW | **TCGGACACCTTGCCTGAAGT** |
| PGRB_RV | **CAGGGCCGAGGGAAGAGTAG** |
| REEP2_FW | **GGGTGCTGTCAGAGAAGCTC** |
| REEP2_RV | **TGTCTCCCATGTCATCCTCA** |
| WNT5A_FW | **TGGCTTTGGCCATATTTTTC** |
| WNT5A_RV | **CCGATGTACTGCATGTGGTC** |
| IHH_FW | **CTCGCCTACAAGCAGTTCAG** |
| IHH_RV | **CCTGTGTTCTCCTCGTCCTT** |
| MMP9_FW | **GCGTCTTCCCCTTCACTTTC** |
| MMP9_RV | **ATAGGGTACATGAGCGCCTC** |
| MSX2_FW | **ATATGAGCCCTACCACCTGC** |
| MSX2_RV | **GCTTTTCCAGTTCTGCCTCC** |
| ESR1_FW | **ATGTGCCTGGCTAGAGATCC** |
| ESR1_RV | **CAAACTCCTCTCCCTGCAGA** |
