## Supplementary Files for "Disrupted PGR-B and ESR1 signaling underlies preconceptional defective decidualization linked to severe preeclampsia"

**Figure supplement**

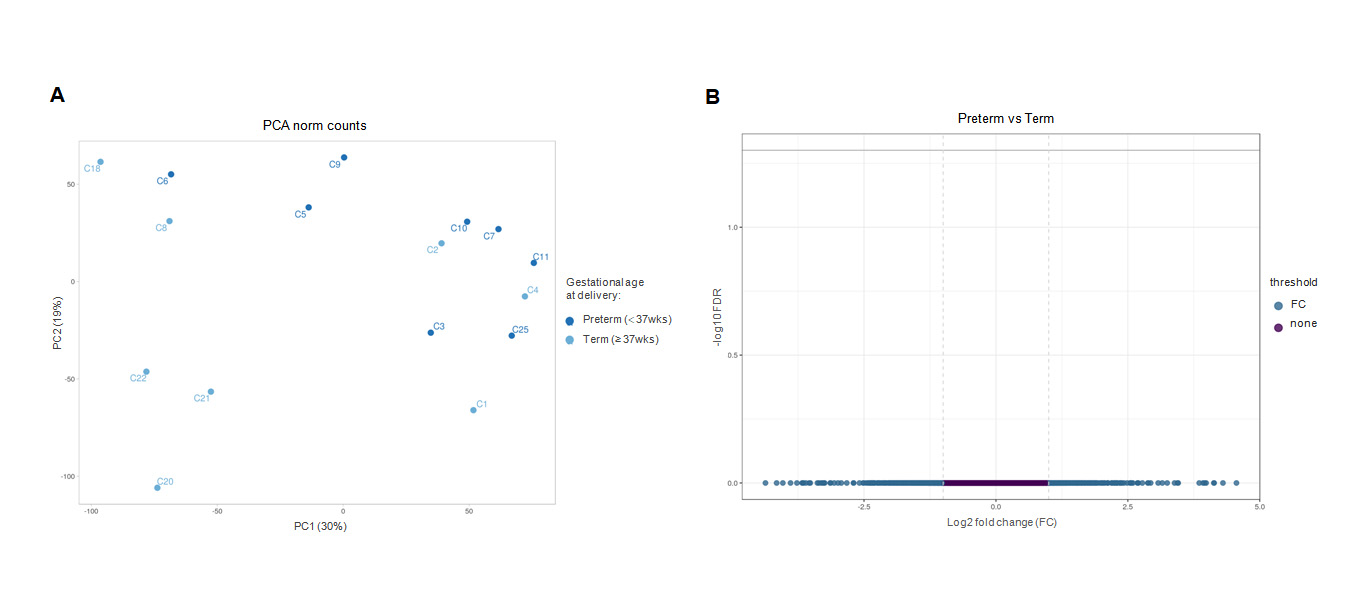

**Figure supplement 1. Transcriptomic analysis based on gestational age at delivery of control samples**. **(A)** PCA based on 18,476 genes after filtering out lowly expressed genes do not demonstrate clustering based on gestational age.  **(B)**  Volcano plot showing the absence of DEGs genes according to gestational age. Legend: none (do not have DEGs genes); fc (high fold-change, but not DEGs genes). Plot based on 728 genes labeled as fc (blue) and 17,748 genes labeled as none (purple).

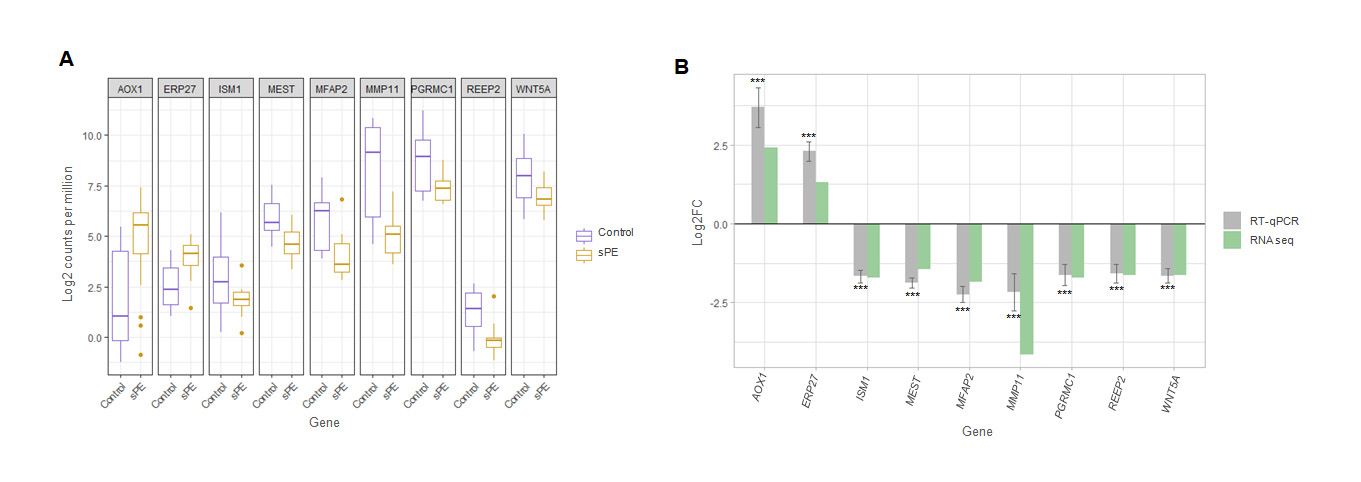

**Figure supplement 2. Validation of RNA-seq results for nine relevant genes. (A)** Boxplot showing the expression patterns of the nine genes obtained in RNA-seq in controls (blue boxes), and sPE (orange boxes). **(B)** RT-qPCR (gray bars) validating the sequencing results (green bars). RT-qPCR values are expressed as mean± SE. *** p≤0.001.

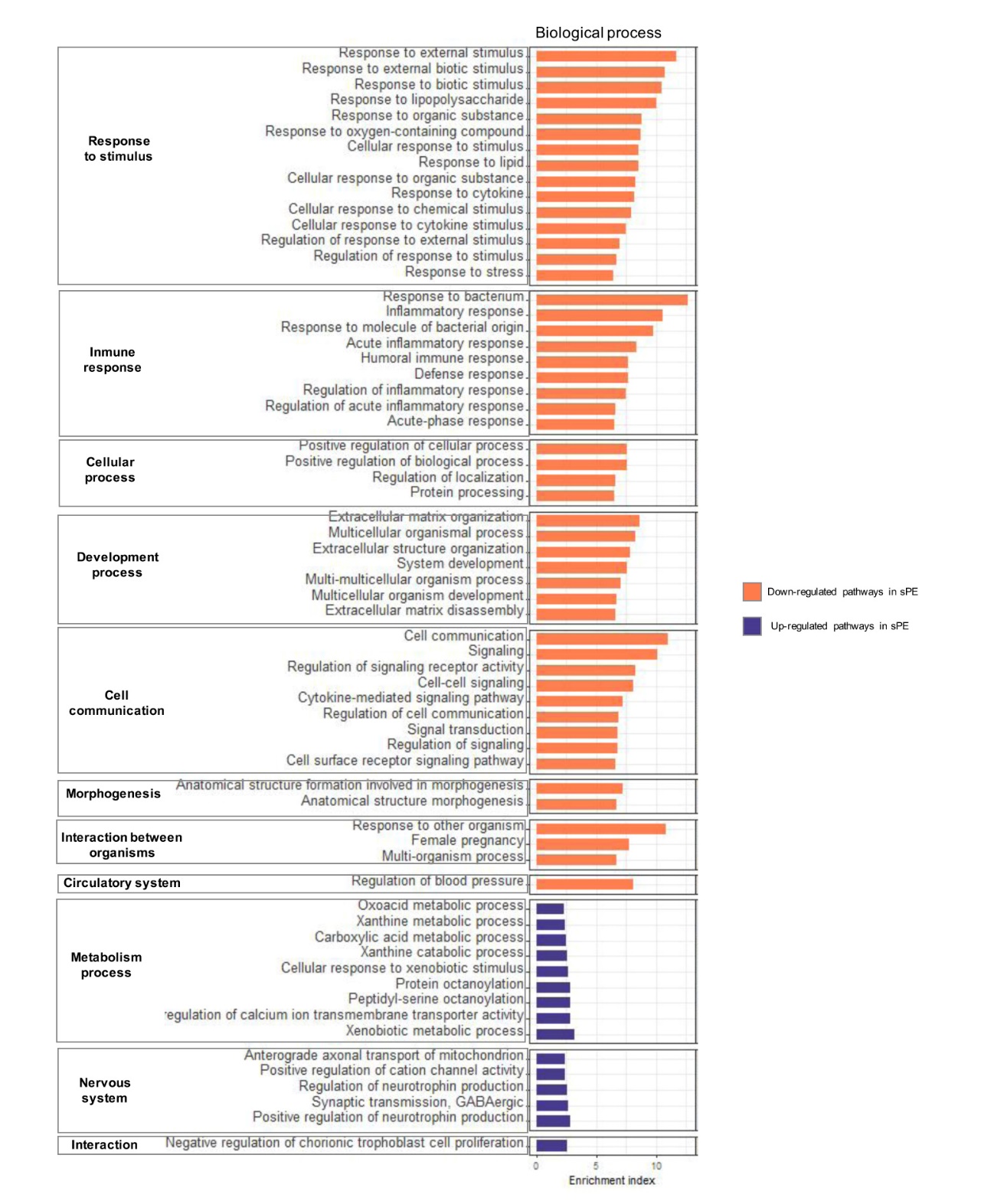

**Figure supplement 3. Functional analysis of fingerprinting encoding decidualization resistance**. **(A)** Gene ontology enrichment analysis of biologic processes. **(B)** Gene ontology enrichment analysis of molecular function. **(C)** Distribution of implicated functions based on the Kyoto of Genes and Genomes pathway database. Enrichment index was calculated as –Log10 (p-value).

**Supplementary files**

**Supplementary file 1.** Maternal and neonatal characteristics of endometrial donors.

**Supplementary file 2.** Differentially expressed genes in sPE vs control cases obtained from Global RNA-seq analysis (859 DEGs).

**Supplementary file 3.** List of genes selected as defective decidualization signature in sPE (166 DEGs).

**Supplementary file 4.** Biological process GO terms enriched in sPE.

**Supplementary file 5.** RT-qPCR primers list.
